# Impact of Antidiabetic Medications on IgG and Plasma Protein N-Glycosylation in Type 2 Diabetes Patients

**DOI:** 10.64898/2026.06.17.26355850

**Authors:** Nikol Mraz, Frano Vučković, Tea Pribić, Anica Radoš Kajić, Tomas Matić, Edita Pape Medvidović, Vilma Kolarić, Dario Rahelić, Gordan Lauc, Tamara Štambuk

**Affiliations:** Genos Glycoscience Research Laboratory, Zagreb, Croatia; Vuk Vrhovac University Clinic for Diabetes, Endocrinology and Metabolic Diseases, Merkur University Hospital, Zagreb, Croatia; Catholic University of Croatia, Faculty of Health Studies, Zagreb, Croatia; Catholic University of Croatia, School of Medicine, Zagreb, Croatia; Josip Juraj Strossmayer University of Osijek School of Medicine, Osijek, Croatia; Department of Biochemistry and Molecular Biology, Faculty of Pharmacy and Biochemistry, University of Zagreb, Zagreb, Croatia

## Abstract

**Introduction:** Diabetes is a growing global health challenge, necessitating effective management strategies. Glycosylation, a highly regulated post-translational protein modification, has emerged as a pivotal factor in diabetes pathophysiology. However, the modulation of protein glycosylation by antidiabetic treatment is still largely unknown. This study explored the longitudinal effects of four distinct antidiabetic therapies – metformin, insulin, sodium-glucose cotransporter-2 (SGLT2) inhibitors, and glucagon-like peptide-1 receptor agonists (GLP-1RA) – on plasma protein and immunoglobulin G (IgG) glycosylation in patients with type 2 diabetes (T2D).

**Research Design and Methods:** Plasma protein and IgG N-glycans were enzymatically released, purified and chromatographically profiled in a cohort of 124 patients, examined at four time points, to assess therapy-induced glycan alterations. Linear mixed models adjusting for covariates and multiple testing (FDR<0.05) were used to investigate the associations between plasma protein and IgG N-glycosylation and antidiabetic therapy.

**Results:** Our findings reveal that metformin, SGLT2 inhibitors, and GLP-1RA induce significant alterations in IgG glycosylation, including the increased core fucosylation and galactosylation, features associated with a reduced inflammatory IgG potential. Notably, IgG monogalactosylation, previously linked to cardioprotective effects in women, was elevated in response to GLP-1RA and SGLT2 inhibitor treatments. Plasma protein glycosylation changes were more limited, with distinct alterations observed for each therapy. Metformin and GLP-1RA similarly reduced certain fucosylated and sialylated glycans, while SGLT2 inhibitors decreased a high-mannose glycan, previously positively associated with diabetes progression. Insulin therapy had a minimal effect on protein glycosylation, with only one plasma glycan significantly altered.

**Conclusions:** Our findings emphasise the importance of protein glycosylation as a dynamic and responsive marker in T2D treatment. The distinct glycan alterations observed in response to metformin, SGLT2 inhibitors, and GLP-1 receptor agonists provide novel insights into the molecular effects of these therapies, potentially contributing to the development of glycan-based biomarkers for personalized diabetes management.

**What is already known on this topic?:**

✓ Protein glycosylation is known to regulate multiple processes related to glucose homeostasis.
✓ Previous studies have shown that the IgG and plasma protein N-glycome (the entire set of N-glycans covalently attached to the IgG and plasma proteins, respectively) can help identify individuals at increased cardiometabolic risk.
✓ Despite most T2D patients receiving antidiabetic treatment, research on its effects on protein glycosylation remains scarce.

**What this study adds?:**

✓ It is the first study to explore the effects of SGLT2 inhibitor and GLP-1RA therapy on protein glycosylation.
✓ The cardioprotective effects of these drug classes were also reflected in IgG glycosylation, demonstrated by increased monogalactosylation.
✓ Metformin, SGLT2 inhibitors, and GLP-1RA induce significant increase in IgG core fucosylation, a feature associated with reduced inflammatory IgG potential, possibly suggesting a shared pathway in suppressing inflammatory response.

**How this study might affect research, practice or policy?:**

✓ The observed alterations in protein glycosylation across different therapeutic groups highlight its dynamic nature in response to metabolic interventions.
✓ Given the role of glycans in modulating immune function and inflammation, these findings suggest that glycan alterations could serve as potential biomarkers for assessing therapeutic response and disease progression in T2D.
✓ Future studies involving protein glycosylation should account for the potential confounding effects of antidiabetic medications to ensure accurate interpretations.

## INTRODUCTION

Diabetes is a complex, multifactorial chronic condition that affects millions of individuals worldwide, posing a significant health challenge and economic burden on global healthcare systems [1]. As the incidence and prevalence of diabetes continue to rise at an alarming rate, there is an urgent need to deepen our understanding of the biological mechanisms underlying its onset and progression. Effective diabetes management is an intricate process, necessitating continuous medical care, pharmacological interventions, and lifestyle modifications to prevent both acute and long-term complications. Among the various treatment strategies available, antidiabetic medications play a crucial role in regulating blood glucose levels and mitigating the risk of diabetes-associated complications.

To provide the better understanding of underlying processes related to diabetes development and its management, an emerging area of research focuses on glycosylation – highly regulated post-translational protein modification. The glycosylation process denotes the enzymatic addition of glycans, complex carbohydrate molecules, to protein backbones. This biochemical process is increasingly recognized for its critical role in shaping the molecular landscape associated with the onset and progression of type 2 diabetes (T2D). By modulating key aspects of protein function, such as structural stability, enzymatic activity, and molecular interactions, glycosylation significantly influences multiple physiological processes essential for glucose homeostasis. Specifically, it plays a regulatory role in glucose sensing, cellular glucose uptake, insulin secretion, and insulin sensitivity [2–4]. Additionally, glycosylation has been implicated in the development of insulin resistance [5] and is closely linked to nutrient influx and metabolic sensing through the hexosamine biosynthetic pathway [6,7].

Our previous research has provided compelling evidence supporting the value of glycan biomarkers in predicting future T2D development, independently of traditional risk factors. Notably, we have shown that baseline differences in plasma protein N-glycome composition can successfully predict future T2D development, identifying at risk individuals up to a decade before diabetes diagnosis [8]. Furthermore, we have revealed that changes in the plasma N-glycome occur gradually, becoming apparent years before the clinical diagnosis of insulin resistance or T2D, with a continuous shift as the disease onset approaches [9]. Given the substantial responsiveness of glycosylation patterns to various environmental and metabolic influences, dynamic changes in glycans could serve as a valuable tool for monitoring the effects of lifestyle modifications. Beneficial interventions, including caloric restriction, increased physical activity, and weight loss, have been shown to induce favourable alterations in protein glycosylation patterns [10–12]. Additionally, plasma protein glycosylation patterns were shown to respond to pharmacological treatments, including widely prescribed medications such as metformin and statins [13]. However, existing studies have largely been cross-sectional, and a comprehensive longitudinal investigation into the effects of medications remains limited.

Recognising that a deeper understanding of these molecular changes could provide novel insights into diabetes prediction and management, we aimed to investigate, for the first time, the longitudinal alterations in plasma protein and immunoglobulin G (IgG) glycosylation following the initiation of antidiabetic therapy in patients with T2D. In our study, we systematically examined the impact of four distinct classes of antidiabetic medications - metformin, sodium-glucose cotransporter-2 (SGLT2) inhibitors, glucagon-like peptide-1 receptor (GLP-1RA) agonists, and insulin - assessing glycosylation changes at four different time points. By characterizing these molecular alterations over time, our research aims to provide insights into the modifications of protein glycosylation by diabetes management, with potential implications for personalised treatment strategies.

## METHODS

### Study population

All subjects participating in this study were recruited at Vuk Vrhovac University Clinic for Diabetes, Endocrinology and Metabolic Diseases. The study included 124 subjects with diabetes, each sampled at up to four timepoints (Table 1). Participants had initiated one of the following antidiabetic treatments: metformin, insulin, an SGLT2 inhibitor, or a GLP-1RA. Diabetes was diagnosed according to the following criteria: fasting plasma glucose ≥7.0 mmol/L and/or a non-fasting plasma glucose level ≥11.1 mmol/L measured at least at two separate time points.

**Table 1.** Overall patient recruitment rate.

|  | **t0** | **t1 (3 months)** | **t2 (6 months)** | **t3 (9-12 months)** |
| --- | --- | --- | --- | --- |
| **Group 1 (metformin)** | 44 | 39 | 32 | 23 |
| **Group 2 (insulin)** | 5 | 4 | 3 | 1 |
| **Group 3 (SGLT2i)** | 50 | 43 | 37 | 33 |
| **Group 4 (GLP-1RA)** | 23 | 18 | 18 | 15 |
| **Total** | **124** | **105** | **90** | **73** |

A very small number of patients were included in the group with newly initiated insulin therapy due to the prioritisation of newer therapeutic options, such as SGLT2 inhibitors and GLP-1RA. Insulin monotherapy is in the Clinic most often initiated in the emergency department for patients with previously undiagnosed, severely uncontrolled diabetes, which significantly limits the pool of eligible participants for the study.

Plasma samples were collected from every patient for the purpose of glycan profiling. Biochemical traits such glycosylated haemoglobin (HbA1c), fasting plasma glucose and insulin, high-sensitivity C-reactive protein (hsCRP), renal function and lipid profile were measured.

This study was approved by the Ethics Committee of the Merkur University Hospital (Approval No. 03/1-7763/1) as a part of the IRI Cardiometabolic project, where routinely collected samples and medical data from regular medical check-up were used. All subjects signed an informed consent, and the study was conducted in accordance with the Declaration of Helsinki. Basic characteristics of the study population at baseline are presented in Table 2.

**Table 2.**
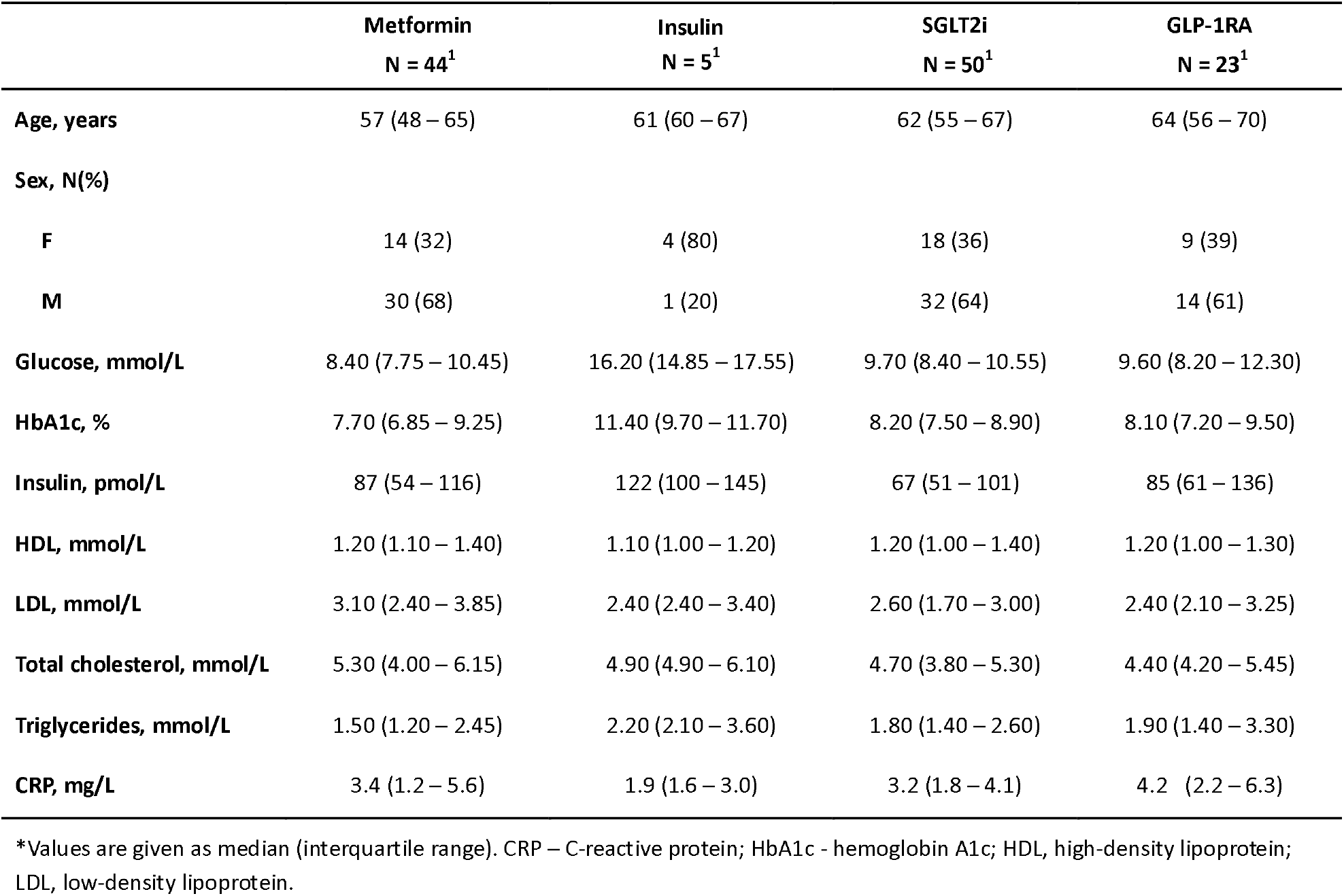
Basic clinical characteristics of study population at baseline*.

|  | **Metformin**<br>**N = 44<sup>1</sup>** | **Insulin**<br>**N = 5<sup>1</sup>** | **SGLT2i**<br>**N = 50<sup>1</sup>** | **GLP-1RA**<br>**N = 23<sup>1</sup>** |
| --- | --- | --- | --- | --- |
| **Age, years** | 57 (48 – 65) | 61 (60 – 67) | 62 (55 – 67) | 64 (56 – 70) |
| **Sex, N(%)** |  |  |  |  |
| **F** | 14 (32) | 4 (80) | 18 (36) | 9 (39) |
| **M** | 30 (68) | 1 (20) | 32 (64) | 14 (61) |
| **Glucose, mmol/L** | 8.40 (7.75 – 10.45) | 16.20 (14.85 – 17.55) | 9.70 (8.40 – 10.55) | 9.60 (8.20 – 12.30) |
| **HbA1c, %** | 7.70 (6.85 – 9.25) | 11.40 (9.70 – 11.70) | 8.20 (7.50 – 8.90) | 8.10 (7.20 – 9.50) |
| **Insulin, pmol/L** | 87 (54 – 116) | 122 (100 – 145) | 67 (51 – 101) | 85 (61 – 136) |
| **HDL, mmol/L** | 1.20 (1.10 – 1.40) | 1.10 (1.00 – 1.20) | 1.20 (1.00 – 1.40) | 1.20 (1.00 – 1.30) |
| **LDL, mmol/L** | 3.10 (2.40 – 3.85) | 2.40 (2.40 – 3.40) | 2.60 (1.70 – 3.00) | 2.40 (2.10 – 3.25) |
| **Total cholesterol, mmol/L** | 5.30 (4.00 – 6.15) | 4.90 (4.90 – 6.10) | 4.70 (3.80 – 5.30) | 4.40 (4.20 – 5.45) |
| **Triglycerides, mmol/L** | 1.50 (1.20 – 2.45) | 2.20 (2.10 – 3.60) | 1.80 (1.40 – 2.60) | 1.90 (1.40 – 3.30) |
| **CRP, mg/L** | 3.4 (1.2 – 5.6) | 1.9 (1.6 – 3.0) | 3.2 (1.8 – 4.1) | 4.2 (2.2 – 6.3) |

\*Values are given as median (interquartile range). CRP – C-reactive protein; HbA1c - hemoglobin A1c; HDL, high-density lipoprotein; LDL, low-density lipoprotein.

### N-glycome measurements

#### Isolation of Immunoglobulin G (IgG) from human plasma

To isolate IgG from a 25 μL of plasma sample, the CIM^®^r-Protein G LLD 0.05 mL Monolithic 96-well Plate (2 µm channels; BIA Separations, Slovenia) was used according to the previously published protocol [14]. Immunoglobulin G was eluted from a protein G plate using 0.1 M formic acid, pH 2.5 (Sigma Aldrich, USA), and neutralized with 1 M ammonium bicarbonate (Acros Organic, USA).

Subsequently, 130 μL of IgG eluate was aliquoted in a 1 mL collection plate (Waters, MA, USA) and dried in a Savant SpeedVac vacuum concentrator (Thermo-Fischer Scientific, USA).

#### Total plasma protein and IgG N-glycan release, fluorescent labelling and HILIC-SPE purification

After drying, IgG was denatured with addition of 30 μL of 1.33% SDS (w/v) (Invitrogen, USA) followed by incubation at 65°C for 10 min. Plasma samples (10 μl) were denatured with the addition of 20 μl of 2% (w/v) SDS and by incubation at 65°C for 10 min. From this point on, the procedure was identical for both IgG and total plasma protein samples. After denaturation, 10 μl of 4% (v/v) Igepal-CA630 (Sigma Aldrich, USA) was added to the samples, followed by glycan release, by adding 1.2 U PNGase F (Promega, USA) and incubating overnight at 37 °C. Released plasma N-glycans were labelled with 2-aminobenzamide (2-AB, Sigma Aldrich, USA). The labelling mixture consisted of 0.48 mg of 2-AB and 1.12 mg of 2-picoline borane (2-PB, Sigma-Aldrich) in 25 μl of dimethyl sulfoxide (DMSO, Sigma-Aldrich) and glacial acetic acid (Merck, Darmstadt, Germany) (7:3, v/v) per sample. The labelling mixture was added to each sample followed by incubation at 65 °C for 2 hours. Excess reagents and proteins were removed from the samples by solid phase extraction by hydrophilic interaction liquid chromatography (HILIC-SPE) using 0.2 μm wwPTFE 96-well membrane filter plates (Pall, NY, USA), as previously described [15]. N-glycans were eluted with water and stored at -20 °C until further analysis.

### N-glycan hydrophilic interaction liquid chromatography analysis

Fluorescently labelled N-glycans were separated by hydrophilic interaction chromatography on Acquity UPLC H-Class instrument (Waters, USA) consisting of a quaternary solvent manager, sample manager and a fluorescence detector set with excitation and emission wavelengths of 250 and 428 nm, respectively. The instrument was under the control of Empower 3 software, build 3.6.1 (Waters, USA). N-glycans were separated on a Waters Glycan Premier BEH amide chromatography column, with 100 mM ammonium formate, pH 4.4, as solvent A and ACN as solvent B. In the case of IgG N-glycans, separation method used linear gradient of 75–62% acetonitrile at flow rate of 0.4 ml/min in a 27-minute analytical run. For plasma protein N-glycans separation method used linear gradient of 70–53% acetonitrile at flow rate of 0.56 ml/min in a 25-minute analytical run. The chromatograms were all separated in the same manner into 24 peaks (GP1-GP24) for IgG N-glycans and 39 peaks (GP1-GP39) for plasma protein N-glycans (Supplementary Figure 1 and 2). The glycan peaks were analysed by their elution positions, and the amount of glycans in each peak was expressed as a percentage of the total integrated area. For IgG glycans, in addition to 24 directly measured glycan traits, 9 derived traits were calculated. For plasma glycans, in addition to 39 directly measured glycan traits, 16 derived traits were calculated. These derived traits average distinct glycosylation features, such as glycan branching, galactosylation, fucosylation, presence of N-acetylglucosamine (GlcNAc), and sialylation (Supplementary Table 1).

### Statistical analysis

To remove experimental variation from the measurements, normalization and batch correction were performed on the raw UPLC glycan data. To make measurements across samples comparable, normalization by total area was performed. Prior to batch correction, normalized glycan measurements were log-transformed due to right-skewness of their distributions and the multiplicative nature of batch effects. Batch correction was performed on log-transformed measurements using the ComBat method (R package sva [16]), where the technical source of variation (which sample was analysed on which plate) was modelled as batch covariate. To correct measurements for experimental noise, estimated batch effects were subtracted from log-transformed measurements.

To assess baseline differences in glycosylation between the four therapeutic groups, a regression model was used. Analyses included glycan measurement as dependent continuous variable, therapeutic group was included as independent variable, with age and sex included as additional covariates. Longitudinal analysis of patient samples through their observation period was performed by implementing a linear mixed effects model, where time was modeled both as a fixed effect and random slope, age and sex were modelled as fixed effects, while individual ID was modeled as a random intercept. Prior to the analyses, glycan variables were all transformed to standard Normal distribution by inverse transformation of ranks to Normality (R package “GenABEL”, function rntransform). Using rank transformed variables makes estimated effects of different glycans comparable, as these will have the same standardized variance. False discovery rate (FDR) was controlled by the Benjamini-Hochberg procedure at the specified level of 0.05. Data was analysed and visualized using R programming language (version 4.3.2) [17].

## RESULTS

N-glycome profiles of both plasma protein and IgG were analysed in a cohort of patients initiating new antidiabetic treatments, across four distinct timepoints. Firstly, differences in glycosylation profiles among patients receiving drugs from four therapeutic groups were assessed. Baseline glycosylation patterns of IgG and total plasma proteins were compared to confirm that any longitudinal changes were driven by therapy. Each therapeutic group was iteratively compared with the other three (Supplementary Table 2). To assess baseline differences in glycosylation between the four therapeutic groups, a regression model was used. Analyses included glycan measurement as dependent continuous variable, therapeutic group was included as independent variable, with age and sex included as additional covariates. The analysis included 132 combinations of tested differences in glycan profiles depending on the introduced antidiabetic therapy (metformin vs insulin vs SGLT2 inhibitors vs GLP-1RA) and no statistically significant differences were observed. These findings confirm that, at baseline, glycosylation patterns did not differ among T2D patient groups, ensuring that any observed longitudinal effects were attributable to the introduced therapies.

### Impact of Metformin Therapy on Protein Glycosylation

To identify the protein glycosylation changes associated with metformin therapy, a linear mixed model was used. We detected substantial changes in the IgG N-glycome, with 7 out of 24 initially measured glycans, and 3 out of 9 calculated derived traits showing significant alterations following the therapy introduction (Table 3). Specifically, we observed an increase in the levels of core fucosylated (CF), digalactosylated (G2) IgG glycans and glycans with bisecting GlcNAc (B) structures (Table 3). In contrast, individual glycan structures without core fucose bearing one or two sialic acids (GP16, GP17, GP21) were significantly decreased, however, total IgG sialylation (S) was not significantly decreased. Figure 1 illustrates the longitudinal changes in IgG glycosylation traits following the introduction of antidiabetic therapy.

**Table 3.**
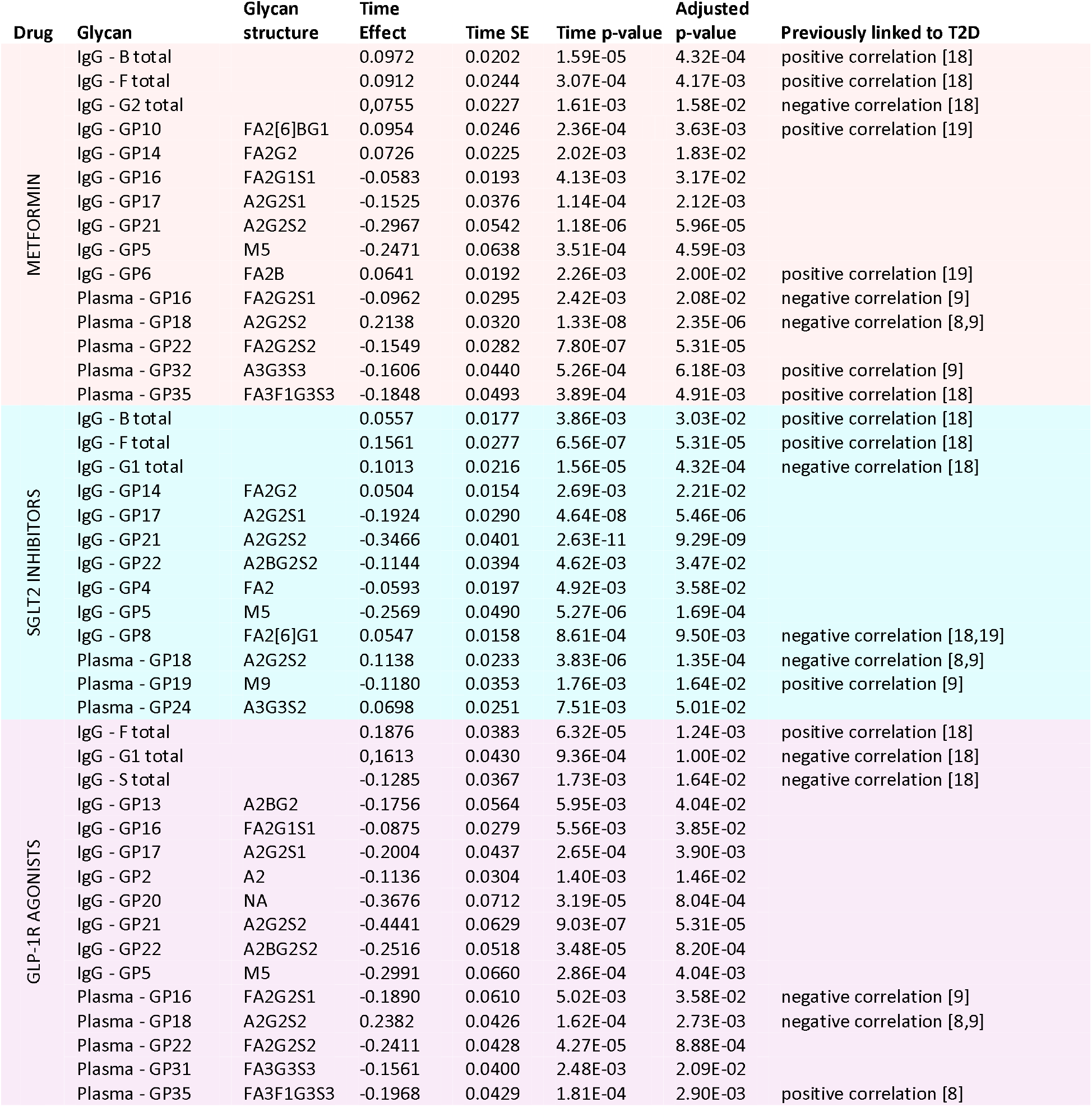
Longitudinal effects of antidiabetic therapy on IgG and total plasma protein N-glycosylation. Longitudinal analysis was performed by implementing a linear mixed effects model, with time as a fixed effect and the individual sample measurement as a random effect. False discovery rate was controlled using Benjamini-Hochberg method at the specified level of 0.05. Only significant results are listed.

| Drug | Glycan | Glycan structure | Time Effect | Time SE | Time p-value | Adjusted p-value | Previously linked to T2D |
| --- | --- | --- | --- | --- | --- | --- | --- |
| METFORMIN | IgG - B total |  | 0.0972 | 0.0202 | 1.59E-05 | 4.32E-04 | positive correlation [18] |
|  | IgG - F total |  | 0.0912 | 0.0244 | 3.07E-04 | 4.17E-03 | positive correlation [18] |
|  | IgG - G2 total |  | 0,0755 | 0.0227 | 1.61E-03 | 1.58E-02 | negative correlation [18] |
|  | IgG - GP10 | FA2[6]BG1 | 0.0954 | 0.0246 | 2.36E-04 | 3.63E-03 | positive correlation [19] |
|  | IgG - GP14 | FA2G2 | 0.0726 | 0.0225 | 2.02E-03 | 1.83E-02 |  |
|  | IgG - GP16 | FA2G1S1 | -0.0583 | 0.0193 | 4.13E-03 | 3.17E-02 |  |
|  | IgG - GP17 | A2G2S1 | -0.1525 | 0.0376 | 1.14E-04 | 2.12E-03 |  |
|  | IgG - GP21 | A2G2S2 | -0.2967 | 0.0542 | 1.18E-06 | 5.96E-05 |  |
|  | IgG - GP5 | M5 | -0.2471 | 0.0638 | 3.51E-04 | 4.59E-03 |  |
|  | IgG - GP6 | FA2B | 0.0641 | 0.0192 | 2.26E-03 | 2.00E-02 | positive correlation [19] |
|  | Plasma - GP16 | FA2G2S1 | -0.0962 | 0.0295 | 2.42E-03 | 2.08E-02 | negative correlation [9] |
|  | Plasma - GP18 | A2G2S2 | 0.2138 | 0.0320 | 1.33E-08 | 2.35E-06 | negative correlation [8,9] |
|  | Plasma - GP22 | FA2G2S2 | -0.1549 | 0.0282 | 7.80E-07 | 5.31E-05 |  |
|  | Plasma - GP32 | A3G3S3 | -0.1606 | 0.0440 | 5.26E-04 | 6.18E-03 | positive correlation [9] |
|  | Plasma - GP35 | FA3F1G3S3 | -0.1848 | 0.0493 | 3.89E-04 | 4.91E-03 | positive correlation [18] |
| SGLT2 INHIBITORS | IgG - B total |  | 0.0557 | 0.0177 | 3.86E-03 | 3.03E-02 | positive correlation [18] |
|  | IgG - F total |  | 0.1561 | 0.0277 | 6.56E-07 | 5.31E-05 | positive correlation [18] |
|  | IgG - G1 total |  | 0.1013 | 0.0216 | 1.56E-05 | 4.32E-04 | negative correlation [18] |
|  | IgG - GP14 | FA2G2 | 0.0504 | 0.0154 | 2.69E-03 | 2.21E-02 |  |
|  | IgG - GP17 | A2G2S1 | -0.1924 | 0.0290 | 4.64E-08 | 5.46E-06 |  |
|  | IgG - GP21 | A2G2S2 | -0.3466 | 0.0401 | 2.63E-11 | 9.29E-09 |  |
|  | IgG - GP22 | A2BG2S2 | -0.1144 | 0.0394 | 4.62E-03 | 3.47E-02 |  |
|  | IgG - GP4 | FA2 | -0.0593 | 0.0197 | 4.92E-03 | 3.58E-02 |  |
|  | IgG - GP5 | M5 | -0.2569 | 0.0490 | 5.27E-06 | 1.69E-04 |  |
|  | IgG - GP8 | FA2[6]G1 | 0.0547 | 0.0158 | 8.61E-04 | 9.50E-03 | negative correlation [18,19] |
|  | Plasma - GP18 | A2G2S2 | 0.1138 | 0.0233 | 3.83E-06 | 1.35E-04 | negative correlation [8,9] |
|  | Plasma - GP19 | M9 | -0.1180 | 0.0353 | 1.76E-03 | 1.64E-02 | positive correlation [9] |
|  | Plasma - GP24 | A3G3S2 | 0.0698 | 0.0251 | 7.51E-03 | 5.01E-02 |  |
| GLP-1R AGONISTS | IgG - F total |  | 0.1876 | 0.0383 | 6.32E-05 | 1.24E-03 | positive correlation [18] |
|  | IgG - G1 total |  | 0,1613 | 0.0430 | 9.36E-04 | 1.00E-02 | negative correlation [18] |
|  | IgG - S total |  | -0.1285 | 0.0367 | 1.73E-03 | 1.64E-02 | negative correlation [18] |
|  | IgG - GP13 | A2BG2 | -0.1756 | 0.0564 | 5.95E-03 | 4.04E-02 |  |
|  | IgG - GP16 | FA2G1S1 | -0.0875 | 0.0279 | 5.56E-03 | 3.85E-02 |  |
|  | IgG - GP17 | A2G2S1 | -0.2004 | 0.0437 | 2.65E-04 | 3.90E-03 |  |
|  | IgG - GP2 | A2 | -0.1136 | 0.0304 | 1.40E-03 | 1.46E-02 |  |
|  | IgG - GP20 | NA | -0.3676 | 0.0712 | 3.19E-05 | 8.04E-04 |  |
|  | IgG - GP21 | A2G2S2 | -0.4441 | 0.0629 | 9.03E-07 | 5.31E-05 |  |
|  | IgG - GP22 | A2BG2S2 | -0.2516 | 0.0518 | 3.48E-05 | 8.20E-04 |  |
|  | IgG - GP5 | M5 | -0.2991 | 0.0660 | 2.86E-04 | 4.04E-03 |  |
|  | Plasma - GP16 | FA2G2S1 | -0.1890 | 0.0610 | 5.02E-03 | 3.58E-02 | negative correlation [9] |
|  | Plasma - GP18 | A2G2S2 | 0.2382 | 0.0426 | 1.62E-04 | 2.73E-03 | negative correlation [8,9] |
|  | Plasma - GP22 | FA2G2S2 | -0.2411 | 0.0428 | 4.27E-05 | 8.88E-04 |  |
|  | Plasma - GP31 | FA3G3S3 | -0.1561 | 0.0400 | 2.48E-03 | 2.09E-02 |  |
|  | Plasma - GP35 | FA3F1G3S3 | -0.1968 | 0.0429 | 1.81E-04 | 2.90E-03 | positive correlation [8] |

**Figure 1.**
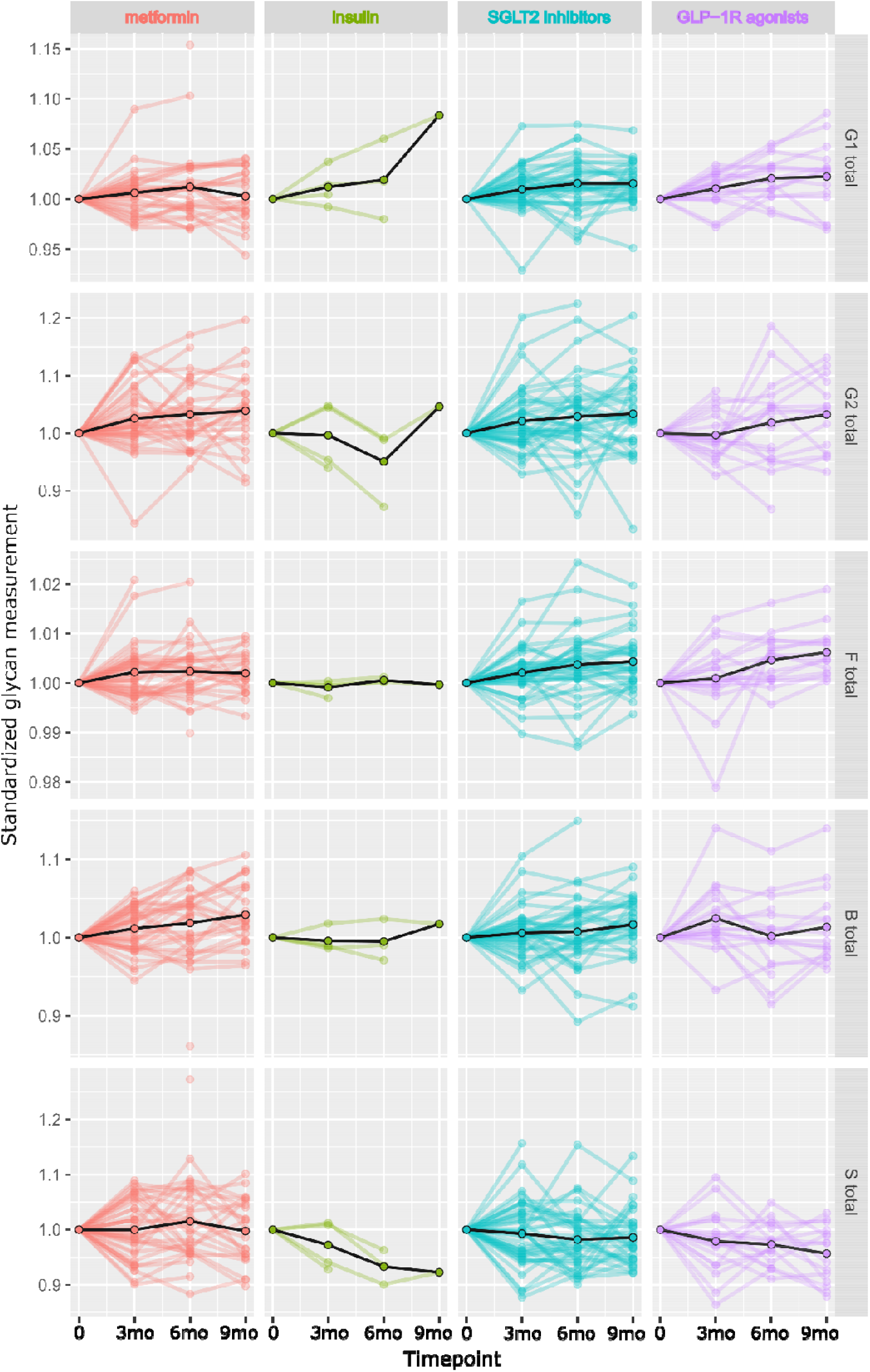
Longitudinal alterations in IgG N-glycome following the introduction of antidiabetic therapy.

In the case of plasma protein glycome, we observed decreased levels of four distinct glycans (GP16 - FA2G2S1, GP22 - FA2G2S2. GP32 - A3G3S3 and GP35 - FA3F1G3S3) and an increase in A2G2S2 glycan (GP18) following the introduction of metformin (Table 3). Most of these glycans have previously been associated with T2D. Interestingly, none of the 16 tested derived plasma glycan traits showed significant change in response to metformin. Figure 2 illustrates the significant longitudinal changes in plasma protein glycans following the introduction of antidiabetic therapy.

**Figure 2.**
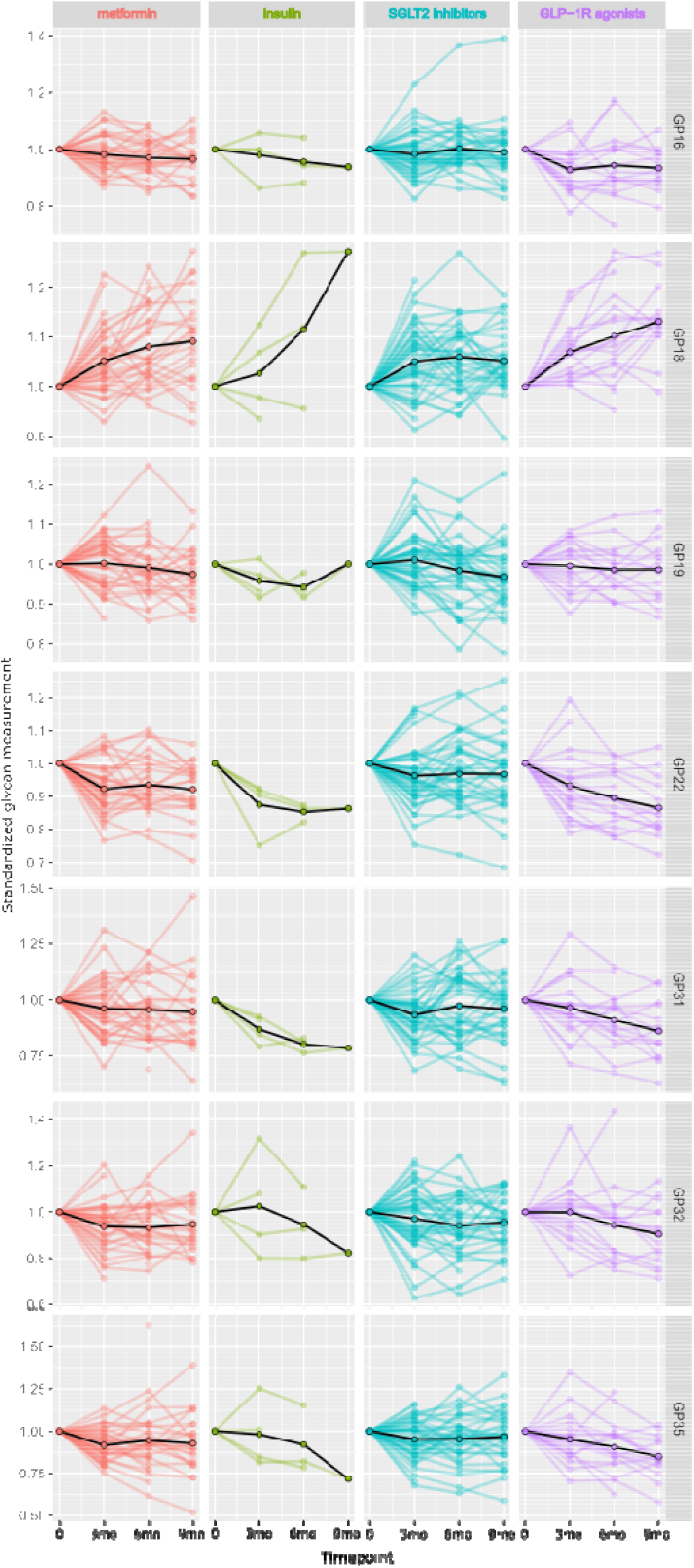
Significant longitudinal alterations in plasma protein N-glycome following the introduction of antidiabetic therapy.

### Impact of Insulin Therapy on Protein Glycosylation

To identify the protein glycosylation changes associated with insulin therapy. the same linear mixed model was used. However, the analysis was burdened by a very small number of patients included in the group with newly introduced insulin therapy. Thus, IgG glycome did not exhibit any significant alterations in response to insulin therapy (Figure 1). However, a single plasma protein N-glycan was significantly decreased – fucosylated and fully sialylated trianntenary structure FA3G3S3 (GP31; effect=-0.538; p-adjusted=0.0358) (Fig. 2), which hasn’t been associated with T2D in previous research.

### Impact of SGLT2 Inhibitor Therapy on Protein Glycosylation

Significant alterations were identified in the IgG N-glycome composition following the introduction of the SGLT2 inhibitor therapy. Specifically, 7 out of 24 initially measured IgG glycans alongside 3 out of 9 tested derived traits showed notable changes following this treatment in T2D patients (Table 3). Similarly to the metformin therapy, we observed an increase in core fucosylated (CF) and bisected (B) glycan structures, however here monogalactosylated (G1) instead of digalactosylated (G2) glycans were increased (Table 3). Conversely, the most abundant agalactosylated glycan (GP4 – FA2) and various individual glycan structures lacking core fucose and carrying one or two sialic acids (GP17, GP22, GP22) exhibited significant reductions. Figure 1 illustrates the longitudinal changes in IgG glycosylation traits following the introduction of antidiabetic therapy.

The plasma protein glycome responded to SGLT2 inhibitor therapy with a decline in one specific high-mannose glycan (GP19 – M9) and concomitant increase in disialylated A2G2S2 (GP18) and A3G3S2 (GP24) glycans (Table 3). None of the 16 analysed derived plasma glycan traits demonstrated significant changes in response to SGLT2 inhibitors. Notably, several significantly altered IgG and plasma N glycans have been previously linked to T2D. Figure 2 illustrates the significant longitudinal changes in plasma protein glycans following the introduction of antidiabetic therapy.

### Impact of GLP-1RA Therapy on Protein Glycosylation

Lastly, we detected substantial changes in the IgG and plasma N-glycome following the introduction of GLP-1RA therapy in T2D patients. In the case of IgG N-glycome, 8 out of 24 initially measured glycans, and 3 out of 9 calculated derived traits showed significant alterations (Table 3). Specifically, we observed an increase in the levels of core fucosylated (CF) and monogalactosylated (G1) IgG glycans (same as in SGLT2 inhibitor response) and a decrease in IgG sialylation (S), which wasn’t observed in the previous antidiabetics’ groups. Figure 1 illustrates the longitudinal changes in IgG glycosylation traits following the introduction of antidiabetic therapy.

Plasma protein N-glycome showed significant alterations in 5 out of 39 initially measured glycans; however, none of the calculated derived glycan traits exhibited significant changes in response to GLP-1RA. Specifically, levels of several fucosylated and sialylated plasma glycans (FA2G2S1 (GP16), FA2G2S2(GP22), FA3G3S3 (GP31) and FA3F1G3S3 (GP32)) were significantly decreased whereas the level of A2G2S2 (GP18) glycan was increased (Table 3). Very similar effect was observed in the metformin group as well, and majority of these glycans have previously been associated with T2D. Figure 2 illustrates the significant longitudinal changes in plasma protein glycans following the introduction of antidiabetic therapy.

## DISCUSSION

In this study, we explored the longitudinal effects of different antidiabetic therapies on plasma protein and IgG glycosylation, including metformin, insulin, SGLT2 inhibitors, and GLP-1RA. Our findings suggest that protein glycosylation undergoes significant alterations in response to specific antidiabetic treatments, underscoring the potential role of glycan profiling in understanding disease progression and therapeutic efficacy in T2D.

Our results showed significant changes in the composition of IgG N-glycome associated with the introduction of antidiabetic therapy. Namely, use of GLP-1RA and SGLT2 inhibitors induced an increase in IgG monogalactosylation – a glycan feature that was previously associated with cardioprotective effects, and marked as a good predictor of major adverse cardiovascular events (MACE) in women [8,18,20]. Both GLP-1RA and SGLT2 inhibitors are relatively new classes of antihyperglycemic drugs, that were also shown to reduce cardiovascular risk [21]. Specifically, these drug classes similarly reduced MACE, with GLP-1RA showing a 12% reduction and SGLT2 inhibitors an 11% risk reduction [22]. However, these benefits were observed only in patients with established atherosclerotic cardiovascular disease. Our findings suggest that their cardioprotective effect is also reflected in IgG glycosylation, specifically through increased monogalactosylation, within the first several months of treatment. While the exact mechanism of their cardioprotective effect is still not elucidated, currently it is largely attributed to metabolic improvements, including better weight management, glycaemic control, and reduced inflammation. Furthermore, a consistent increase in IgG fucosylation has been observed in most therapeutic groups, except for insulin, likely due to the small number of subjects in this group. Increased core fucosylation was systematically associated with a decrease in the proinflammatory potential of IgG, since the absence of core fucose massively increases the affinity of IgG for FcγIII receptors (up to 50-fold), leading to significantly enhanced antibody-dependent cellular cytotoxicity by NK cells [23]. Use of metformin has, however, lead to an increase in IgG digalactosylation, which has been shown to mediate both pro- and anti-inflammatory IgG effector functions [24,25]. Interestingly, a growing body of research is showing that metformin, in addition to the aforementioned metabolic benefits also has a direct anti-inflammatory action – it suppresses inflammatory response by inhibition of nuclear factor κB via AMP-activated protein kinase-dependent and independent pathways [26]. Lastly, the use of GLP-1RA was associated with a decrease in IgG sialylation, whereas therapy with metformin and SGLT2 inhibitors increased levels of bisecting GlcNAc. Such changes in IgG glycosylation are usually associated with increased proinflammatory IgG potential [27,28]. We have also revealed that majority of the observed changes in IgG glycosylation in response to antihyperglycemic treatment move in the beneficial direction, i.e., are opposite to those observed during the T2D development and onset. Overall, the observed shifts in IgG glycosylation patterns suggest that these therapies exert overlapping yet distinct effects on IgG effector functions, affecting not only its immunological but also metabolic roles. There is a growing body of evidence that IgG might also have a strong role in metabolic regulation, supported by a recent study which highlighted its unrecognised role in the induction of metabolic dysfunction [29]. The study revealed that IgG accumulated in adipose tissue in obesity, causing macrophage infiltration and adipose inflammation, and this accumulated IgG obstructs binding of insulin to its receptor, promoting insulin resistance. It has also been shown that IgG glycosylation plays a pivotal role in the induction of obesity-related insulin resistance in the skeletal muscle [5]. The proposed mechanism involves a significant reduction in skeletal muscle insulin delivery, triggered by hyposialylated IgG.

This specific IgG glycoform exhibits enhanced binding affinity to endothelial FcγRIIB, leading to decreased insulin transcytosis. In contrast, sialylated IgG does not impair endothelial insulin transcytosis or its delivery to skeletal muscle cells, thereby preserving insulin-induced cell-surface GLUT4 expression, promoting glucose uptake, and maintaining insulin sensitivity. Overall, these findings suggest that the changes in IgG glycosylation induced by antidiabetic therapies, contribute to their cardioprotective effects and potential modulation of insulin sensitivity through distinct mechanisms that involve both immunological and metabolic pathways that warrants further research.

Contrary to IgG, changes in plasma protein glycosylation were more limited, where only distinct glycans responded to the antidiabetic treatment. Specifically, metformin and GLP-1RA both showed a similar effect, causing a decreased levels of several bi- and tri-antennary, fucosylated and sialylated plasma glycans, majority of which have previously been linked to T2D, which further supports the biological relevance of these findings. The direction of glycosylation changes upon the therapy introduction is favourable for vast majority of structures and the observed similarities may point to shared underlying mechanisms between these two therapies, involving improved metabolic control and reduced systemic inflammation. SGLT2 inhibitors led to a reduction in the M9 glycan, high-mannose structure previously found to be elevated during the development of both T1D [30] and T2D [9]. This glycan structure is mostly originating from apolipoprotein B-100 and complement component C3, both with known links to diabetes development [31,32]. Interestingly, all three classes of oral antidiabetics led to an increase in A2G2S2 glycan, which has been negatively associated with diabetes risk and development across several studies [8,9]. Surprisingly, no significant changes in derived plasma glycan traits were observed after the introduction of antidiabetics. The reasons for the lack of response in the composite measures of plasma glycome glycosylation can be multiple: (i) there are large and conflicting inter-individual differences in the direction of change in plasma glycosylation in response to therapy, diminishing the average group effect. (ii) the examined population is not of sufficient size to observe less pronounced changes; (iii) plasma glycome is a complex mixture of N-glycans originating from many different plasma glycoproteins, which may respond differently to the introduction of antidiabetic therapy, masking the induced changes on less abundant glycoproteins. However, there were some indications of the most pronounced changes: metformin therapy led to a nominally significant decrease in core fucosylation, GLP-1RA led to an increase in digalactosylation whereas SGLT2 inhibitors induced a decrease in high-mannose structures on plasma proteins. The use of metformin was previously associated with a decrease in fucosylation of di-, tri- and tetraantennary glycans, irrespective of the presence of sialylation [13], which further corroborates our nominally significant finding here.

Lastly, insulin therapy resulted in minimal changes to the IgG glycome, possibly due to lack of statistical power in this group. Interestingly, a previous cross-sectional study revealed no significant association between insulin use and the plasma protein N-glycome despite good power (with 919 patients on insulin therapy) [13]. In our longitudinal design, the only significant alteration observed was a decrease in a core fucosylated and fully sialylated trianntenary plasma glycan (FA3G3S3), which has not been previously associated with T2D. Based on the literature sources [33], the predominant source of this glycan structure is vitronectin. It is a glycoprotein involved in cell adhesion, with proliferation-mediating properties, and it has also been implicated in various aspects of diabetes complications development, including proliferative diabetic retinopathy [34] and vascular complications [35]. The levels of vitronectin were found to be elevated in patients with diabetes, however, at this point we cannot speculate if the glycosylation change observed influences its function. Nonetheless, its dysregulation may contribute to diabetic complications, especially those related to the cardiovascular system and tissue repair. Overall, the limited glycosylation changes in response to insulin may reflect the heterogeneity of insulin-treated patients, often characterized by more advanced disease stages and distinct metabolic profiles, or insulin, as a naturally occurring hormone, does not substantially influence glycosylation machinery.

Altogether, the observed modifications in glycosylation profiles across different therapeutic groups highlight the dynamic nature of protein glycosylation in response to metabolic interventions. Given the role of glycans in modulating immune function and inflammation, these findings suggest that glycan alterations could serve as potential biomarkers for assessing therapeutic response and disease progression in T2D. Furthermore, the responsiveness of glycosylation to pharmaceutical interventions aligns with previous evidence suggesting that lifestyle changes, such as weight loss [12,36] and physical activity [11,37], can similarly influence IgG and plasma protein glycan composition.

Despite these promising insights, several limitations should be considered. The relatively small sample size, particularly in the insulin-treated group, may have limited our ability to detect significant glycosylation changes in this subset. Additionally, while our study provides valuable longitudinal data, further research is needed to establish the mechanistic links between glycosylation changes and immuno-metabolic improvements in response to therapy. Future investigations should focus on expanding cohort sizes, explore glycoproteomic approaches (to circumvent the limitations of released glycan analysis from complex protein mixtures, such as blood plasma), and integrating glycomics with other omics approaches to gain a more comprehensive understanding of the molecular effects of antidiabetic treatments.

In conclusion, our findings emphasise the importance of protein glycosylation as a dynamic and responsive marker in T2D treatment. The distinct glycan alterations observed in response to metformin, SGLT2 inhibitors, and GLP-1 receptor agonists provide novel insights into the molecular effects of these therapies, potentially contributing to the development of glycan-based biomarkers for personalized diabetes management. Finally, the use of antidiabetic drugs should also be considered as a potential confounder when designing studies involving glycosylation.

## Data Availability

All data produced in the present study are available upon reasonable request to the authors.

## Acknowledgements

The authors would like to thank all the participants of this study and clinical staff of the Vuk Vrhovac University Clinic for Diabetes, Endocrinology and Metabolic Diseases in Zagreb.

## Contributors

NM and TP performed glycomic analyses and reviewed/edited the manuscript. FV performed statistical analyses and reviewed/edited the manuscript. ARK, TM, EPM and VK recruited patients and reviewed/edited the manuscript. DR contributed to the design and coordination of the study, recruited patients, and reviewed/edited the manuscript. GL contributed to the design and coordination of the study, and reviewed/edited the manuscript. TŠ contributed to the design and coordination of the study, interpreted results and wrote the manuscript. All authors approved the final version of the manuscript. TŠ is the guarantor of this work and, as such, had full access to all the data in the study and takes responsibility for the integrity of the data and the accuracy of the data analysis.

## Data availability statement

Data are available on reasonable request. The datasets generated during and/or analysed during the current study are not publicly available. The raw data are subject to ‘Special Categories of Personal Data (Sensitive Data)’ (GDPR, Article 9), therefore raw data sharing is not in line with the privacy principles. Also, the information provided to the participants in the study states that the individual data are only accessible to the researchers, the ethical review board and (local) authorities. The informed consent given by the participants is therefore not sufficient for open access publication of indirectly identifiable data. Datasets are available from the corresponding author on reasonable request.

