## Supplemental files for "Impact of Antidiabetic Medications on IgG and Plasma Protein N-Glycosylation in Type 2 Diabetes Patients"

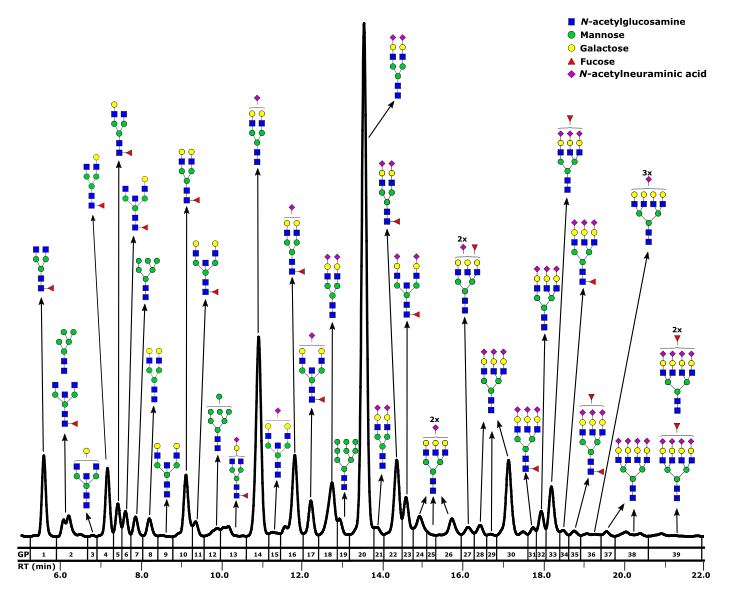

**Supplementary Figure 1** Representative HILIC-UPLC-FLR chromatogram of the total plasma protein N-glycome, with graphic representation of the most abundant glycan structures corresponding to each glycan peak (GP). RT – retention time.

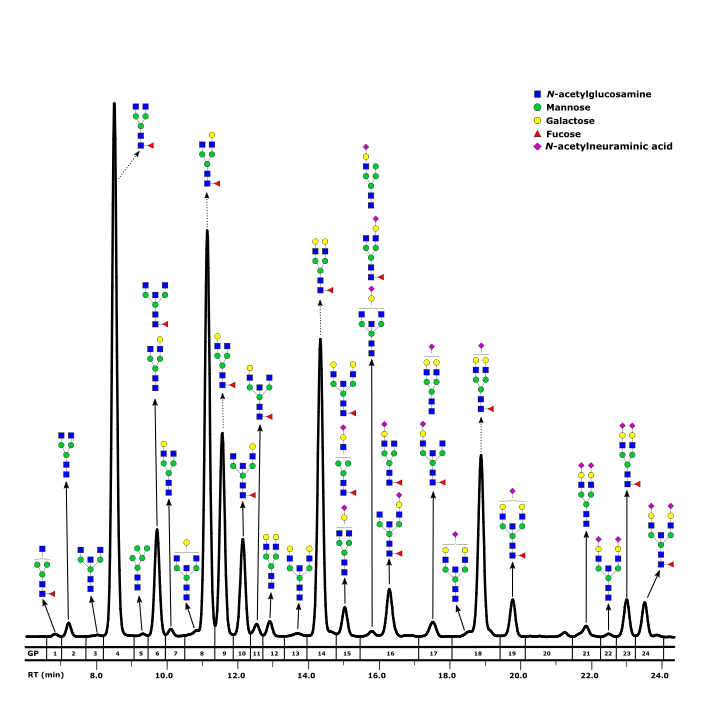

**Supplementary Figure 2** Representative HILIC-UPLC-FLR chromatogram of immunoglobulin G N-glycome, with graphic representation of glycan structures corresponding to each glycan peak (GP). RT – retention time.

**Supplementary Table 1** Derived glycan traits calculated from initial glycan peaks.

| **Derived plasma glycan traits** | **Description** | **Formula** | **Example of a glycan structure** |
| --- | --- | --- | --- |
| LB | Proportion of mono- and biantennary structures in the total plasma N-glycome | GP1+0.5xGP2+GP3+GP4+GP5+GP6+GP8+GP9+GP10+GP11+0.5xGP12+GP13+GP14+GP15+GP16+GP17+GP18+GP20+GP21+GP22+GP23 | 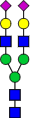 |
| HB | Proportion of tri- and tetraantennary structures in the total plasma N-glycome | GP24+GP25+GP26+GP27+GP28+GP29+GP30+GP31+GP32+GP33+GP34+GP35+GP36+GP37+GP38+GP39 | 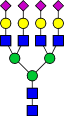 |
| G0 | Proportion of agalactosylated structures in the total plasma N-glycome | GP1+0.5xGP2 | 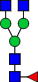 |
| G1 | Proportion of monogalactosylated structures in the total plasma N-glycome | GP3+GP4+GP5+GP6+GP13 | 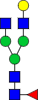 |
| G2 | Proportion of digalactosylated structures in the total plasma N-glycome | GP8+GP9+GP10+GP11+0.5xGP12+GP14+GP15+GP16+GP17+GP18+GP20+GP21+GP22+GP23 | 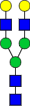 |
| G3 | Proportion of trigalactosylated structures in the total plasma N-glycome | GP24+GP25+GP26+GP27+GP28+GP29+GP30+GP31+GP32+GP33+GP34+GP35 | 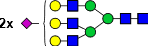 |
| G4 | Proportion of tetragalactosylated structures in the total plasma N-glycome | GP36+GP37+GP38+GP39 | 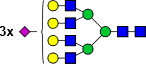 |
| S0 | Proportion of asialylated structures in the total plasma N-glycome | GP1+0.5xGP2+GP3+GP4+GP5+GP6+GP8+GP9+GP10+GP11 | 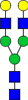 |
| S1 | Proportion of monosialylated structures in the total plasma N-glycome | 0.5xGP12+GP13+GP14+GP15+GP16+GP17 | 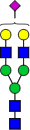 |
| S2 | Proportion of disialylated structures in the total plasma N-glycome | GP18+GP20+GP21+GP22+GP23+GP24+GP25+GP26+GP27 | 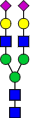 |
| S3 | Proportion of trisialylated structures in the total plasma N-glycome | GP28+GP29+GP30+GP31+GP32+GP33+GP34+GP35+GP36 | 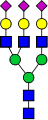 |
| S4 | Proportion of tetrasialylated structures in the total plasma N-glycome | GP37+GP38+GP39 | 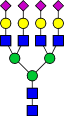 |
| B | Proportion of structures containing bisecting GlcNAc in the total plasma N-glycome | 0.5xGP2+GP3+GP6+GP9+GP11+GP15+GP17+GP23 | 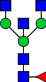 |
| HM | Proportion of high mannose structures in the total plasma N-glycome | 0.5xGP2+GP7+0.5xGP12+GP19 | 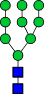 |
| CF | Proportion of structures containing core fucose in the total plasma N-glycome | GP1+0.5xGP2+GP4+GP5+GP6+GP10+GP11+GP13+GP16+GP17+GP22+GP23+GP31+GP34+GP35 | 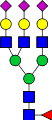 |
| AF | Proportion of structures containing antennary fucose in the total plasma N-glycome | GP27+GP33+GP35+GP39 | 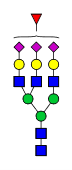 |
| **Derived IgG glycan traits** | **Description** | **Formula** | **Example of a glycan structure** |
| G0 | Proportion of agalactosylated structures in the total IgG N-glycome | GP1+GP2+GP3+GP4+GP5+GP6 | 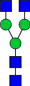 |
| G1 | Proportion of monogalactosylated structures in the total IgG N-glycome | GP7+GP8+GP9+GP10+GP11+GP16 | 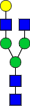 |
| G2 | Proportion of digalactosylated structures in the total IgG N-glycome | GP12+GP13+GP14+GP15+GP17+GP18+GP19+GP21+GP22+GP23+GP24 | 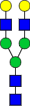 |
| S0 | Proportion of asialylated structures in the total IgG N-glycome | GP1+GP2+GP3+GP4+GP5+GP6+GP7+GP8+GP9+GP10+GP11+GP12+ GP13+GP14+GP15 | 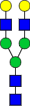 |
| S1 | Proportion of monosialylated structures in the total IgG N-glycome | GP16+GP17+GP18+GP19 | 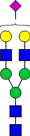 |
| S2 | Proportion of disialylated structures in the total IgG N-glycome | GP21+GP22+GP23+GP24 | 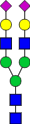 |
| B | Proportion of structures containing bisecting GlcNAc in the total IgG N-glycome | GP3+GP6+GP10+GP11+GP13+GP15+GP19+GP22+GP24 | 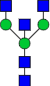 |
| HM | Proportion of high mannose structures in the total IgG N-glycome | GP5 | 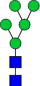 |
| CF | Proportion of structures containing core fucose in the total IgG N-glycome | GP1+GP4+GP6+GP8+GP9+GP10+GP11+GP14+GP15+GP16+GP18+ GP19+GP23+GP24 | 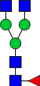 |

GlcNAc – *N*-acetylglucosamine

**Supplementary Table 2** Comparison of IgG and plasma protein glycosylation patterns at baseline (at the introduction of antidiabetic medication) between different therapeutic groups. Analysis was performed by implementing a linear mixed effects model, whereas false discovery rate was controlled using Benjamini-Hochberg method at the specified level of 0.05.

| **Antidiabetic medication comparison** | **Derived glycan trait** | **Effect (β coef.)** | **Standard error** | **p-value** | **Adjusted p-value** |
| --- | --- | --- | --- | --- | --- |
| Metformin - Insulin | IgG - B | 0.9121 | 0.4161 | 0.0258 | 0.3213 |
| Metformin - Insulin | IgG - F | -0.1060 | 0.5042 | 0.8264 | 0.9031 |
| Metformin - Insulin | IgG - G0 | -0.4381 | 0.4182 | 0.2772 | 0.6312 |
| Metformin - Insulin | IgG - G1 | -0.6893 | 0.4958 | 0.1511 | 0.5382 |
| Metformin - Insulin | IgG - G2 | 0.7433 | 0.4075 | 0.0616 | 0.4537 |
| Metformin - Insulin | IgG - S | 0.5774 | 0.4266 | 0.1620 | 0.5382 |
| Metformin - Insulin | Plasma - AF | -0.6170 | 0.4802 | 0.1840 | 0.5392 |
| Metformin - Insulin | Plasma - B | 0.1193 | 0.4940 | 0.8011 | 0.8889 |
| Metformin - Insulin | Plasma - CF | -0.6809 | 0.4962 | 0.1564 | 0.5382 |
| Metformin - Insulin | Plasma - G0 | -0.7537 | 0.4500 | 0.0852 | 0.4537 |
| Metformin - Insulin | Plasma - G1 | -1.2560 | 0.4685 | 0.0070 | 0.1902 |
| Metformin - Insulin | Plasma - G2 | 0.4061 | 0.4690 | 0.3683 | 0.7203 |
| Metformin - Insulin | Plasma - G3 | 0.4953 | 0.4726 | 0.2770 | 0.6312 |
| Metformin - Insulin | Plasma - G4 | -0.3145 | 0.4790 | 0.4943 | 0.7698 |
| Metformin - Insulin | Plasma - HB | 0.3848 | 0.4711 | 0.3958 | 0.7203 |
| Metformin - Insulin | Plasma - HM | 0.8421 | 0.4919 | 0.0787 | 0.4537 |
| Metformin - Insulin | Plasma - LB | -0.6274 | 0.4577 | 0.1568 | 0.5382 |
| Metformin - Insulin | Plasma - S0 | -0.8204 | 0.4854 | 0.0824 | 0.4537 |
| Metformin - Insulin | Plasma - S1 | 0.9340 | 0.4645 | 0.0401 | 0.4056 |
| Metformin - Insulin | Plasma - S2 | 0.0078 | 0.5051 | 0.9872 | 0.9872 |
| Metformin - Insulin | Plasma - S3 | 0.4262 | 0.4701 | 0.3463 | 0.7193 |
| Metformin - Insulin | Plasma - S4 | -0.4521 | 0.4745 | 0.3226 | 0.6875 |
| Metformin - SGLT2 inhibitors | IgG - B | 0.5179 | 0.1819 | 0.0044 | 0.1427 |
| Metformin - SGLT2 inhibitors | IgG - F | -0.2886 | 0.2044 | 0.1513 | 0.5382 |
| Metformin - SGLT2 inhibitors | IgG - G0 | -0.0035 | 0.1897 | 0.9850 | 0.9872 |
| Metformin - SGLT2 inhibitors | IgG - G1 | -0.1429 | 0.2098 | 0.4869 | 0.7658 |
| Metformin - SGLT2 inhibitors | IgG - G2 | 0.1383 | 0.1884 | 0.4539 | 0.7503 |
| Metformin - SGLT2 inhibitors | IgG - S | -0.0512 | 0.1907 | 0.7840 | 0.8854 |
| Metformin - SGLT2 inhibitors | Plasma - AF | -0.4878 | 0.2035 | 0.0159 | 0.2335 |
| Metformin - SGLT2 inhibitors | Plasma - B | -0.0312 | 0.2101 | 0.8793 | 0.9190 |
| Metformin - SGLT2 inhibitors | Plasma - CF | -0.0931 | 0.2131 | 0.6553 | 0.8356 |
| Metformin - SGLT2 inhibitors | Plasma - G0 | 0.1680 | 0.2007 | 0.3932 | 0.7203 |
| Metformin - SGLT2 inhibitors | Plasma - G1 | 0.1583 | 0.2128 | 0.4478 | 0.7479 |
| Metformin - SGLT2 inhibitors | Plasma - G2 | -0.3252 | 0.2077 | 0.1119 | 0.4772 |
| Metformin - SGLT2 inhibitors | Plasma - G3 | 0.2147 | 0.2026 | 0.2804 | 0.6312 |
| Metformin - SGLT2 inhibitors | Plasma - G4 | -0.3588 | 0.2058 | 0.0772 | 0.4537 |
| Metformin - SGLT2 inhibitors | Plasma - HB | 0.1245 | 0.2042 | 0.5337 | 0.7777 |
| Metformin - SGLT2 inhibitors | Plasma - HM | -0.0725 | 0.2128 | 0.7278 | 0.8837 |
| Metformin - SGLT2 inhibitors | Plasma - LB | -0.1266 | 0.2032 | 0.5248 | 0.7777 |
| Metformin - SGLT2 inhibitors | Plasma - S0 | 0.2263 | 0.2109 | 0.2743 | 0.6312 |
| Metformin - SGLT2 inhibitors | Plasma - S1 | -0.2822 | 0.2100 | 0.1717 | 0.5382 |
| Metformin - SGLT2 inhibitors | Plasma - S2 | -0.2747 | 0.2123 | 0.1880 | 0.5392 |
| Metformin - SGLT2 inhibitors | Plasma - S3 | 0.1216 | 0.2026 | 0.5400 | 0.7777 |
| Metformin - SGLT2 inhibitors | Plasma - S4 | -0.3867 | 0.2045 | 0.0556 | 0.4537 |
| Metformin - GLP-1R agonists | IgG - B | 0.3548 | 0.2359 | 0.1242 | 0.5159 |
| Metformin - GLP-1R agonists | IgG - F | -0.1648 | 0.2697 | 0.5291 | 0.7777 |
| Metformin - GLP-1R agonists | IgG - G0 | 0.0892 | 0.2323 | 0.6922 | 0.8623 |
| Metformin - GLP-1R agonists | IgG - G1 | -0.0863 | 0.2642 | 0.7364 | 0.8837 |
| Metformin - GLP-1R agonists | IgG - G2 | -0.0717 | 0.2304 | 0.7483 | 0.8854 |
| Metformin - GLP-1R agonists | IgG - S | 0.1341 | 0.2498 | 0.5803 | 0.8081 |
| Metformin - GLP-1R agonists | Plasma - AF | -0.2774 | 0.2657 | 0.2836 | 0.6312 |
| Metformin - GLP-1R agonists | Plasma - B | 0.2947 | 0.2581 | 0.2414 | 0.6209 |
| Metformin - GLP-1R agonists | Plasma - CF | 0.2261 | 0.2671 | 0.3840 | 0.7203 |
| Metformin - GLP-1R agonists | Plasma - G0 | 0.3167 | 0.2474 | 0.1897 | 0.5392 |
| Metformin - GLP-1R agonists | Plasma - G1 | 0.3744 | 0.2531 | 0.1304 | 0.5280 |
| Metformin - GLP-1R agonists | Plasma - G2 | -0.4371 | 0.2562 | 0.0819 | 0.4537 |
| Metformin - GLP-1R agonists | Plasma - G3 | 0.1314 | 0.2522 | 0.5915 | 0.8081 |
| Metformin - GLP-1R agonists | Plasma - G4 | -0.4569 | 0.2553 | 0.0684 | 0.4537 |
| Metformin - GLP-1R agonists | Plasma - HB | 0.0662 | 0.2513 | 0.7858 | 0.8854 |
| Metformin - GLP-1R agonists | Plasma - HM | 0.1271 | 0.2709 | 0.6288 | 0.8356 |
| Metformin - GLP-1R agonists | Plasma - LB | -0.1036 | 0.2508 | 0.6703 | 0.8417 |
| Metformin - GLP-1R agonists | Plasma - S0 | 0.3230 | 0.2583 | 0.1999 | 0.5489 |
| Metformin - GLP-1R agonists | Plasma - S1 | -0.0677 | 0.2615 | 0.7895 | 0.8854 |
| Metformin - GLP-1R agonists | Plasma - S2 | -0.4068 | 0.2587 | 0.1083 | 0.4742 |
| Metformin - GLP-1R agonists | Plasma - S3 | 0.0951 | 0.2520 | 0.6973 | 0.8623 |
| Metformin - GLP-1R agonists | Plasma - S4 | -0.4328 | 0.2579 | 0.0870 | 0.4537 |
| Insulin - SGLT2 inhibitors | IgG - B | -0.3232 | 0.4369 | 0.4436 | 0.7479 |
| Insulin - SGLT2 inhibitors | IgG - F | 0.0658 | 0.4491 | 0.8791 | 0.9190 |
| Insulin - SGLT2 inhibitors | IgG - G0 | 0.3504 | 0.4341 | 0.4034 | 0.7203 |
| Insulin - SGLT2 inhibitors | IgG - G1 | 0.4059 | 0.4825 | 0.3840 | 0.7203 |
| Insulin - SGLT2 inhibitors | IgG - G2 | -0.3215 | 0.4276 | 0.4361 | 0.7479 |
| Insulin - SGLT2 inhibitors | IgG - S | -0.3308 | 0.4484 | 0.4448 | 0.7479 |
| Insulin - SGLT2 inhibitors | Plasma - AF | -0.0890 | 0.4536 | 0.8386 | 0.9056 |
| Insulin - SGLT2 inhibitors | Plasma - B | -0.0308 | 0.4911 | 0.9480 | 0.9720 |
| Insulin - SGLT2 inhibitors | Plasma - CF | 0.5293 | 0.4902 | 0.2648 | 0.6312 |
| Insulin - SGLT2 inhibitors | Plasma - G0 | 0.5165 | 0.4655 | 0.2521 | 0.6283 |
| Insulin - SGLT2 inhibitors | Plasma - G1 | 0.9392 | 0.4750 | 0.0439 | 0.4179 |
| Insulin - SGLT2 inhibitors | Plasma - G2 | -0.5285 | 0.4931 | 0.2683 | 0.6312 |
| Insulin - SGLT2 inhibitors | Plasma - G3 | -0.4922 | 0.4751 | 0.2844 | 0.6312 |
| Insulin - SGLT2 inhibitors | Plasma - G4 | -0.3342 | 0.4945 | 0.4838 | 0.7658 |
| Insulin - SGLT2 inhibitors | Plasma - HB | -0.5333 | 0.4791 | 0.2506 | 0.6283 |
| Insulin - SGLT2 inhibitors | Plasma - HM | -1.1502 | 0.4628 | 0.0121 | 0.2186 |
| Insulin - SGLT2 inhibitors | Plasma - LB | 0.7805 | 0.4603 | 0.0824 | 0.4537 |
| Insulin - SGLT2 inhibitors | Plasma - S0 | 0.6521 | 0.4862 | 0.1674 | 0.5382 |
| Insulin - SGLT2 inhibitors | Plasma - S1 | -0.7863 | 0.4748 | 0.0896 | 0.4537 |
| Insulin - SGLT2 inhibitors | Plasma - S2 | -0.2733 | 0.4898 | 0.5629 | 0.7930 |
| Insulin - SGLT2 inhibitors | Plasma - S3 | -0.5389 | 0.4709 | 0.2376 | 0.6209 |
| Insulin - SGLT2 inhibitors | Plasma - S4 | -0.3116 | 0.4904 | 0.5102 | 0.7725 |
| Insulin - GLP-1R agonists | IgG - B | -0.5366 | 0.4248 | 0.1794 | 0.5382 |
| Insulin - GLP-1R agonists | IgG - F | -0.3109 | 0.5238 | 0.5230 | 0.7777 |
| Insulin - GLP-1R agonists | IgG - G0 | 0.5677 | 0.4325 | 0.1635 | 0.5382 |
| Insulin - GLP-1R agonists | IgG - G1 | 0.0128 | 0.4510 | 0.9755 | 0.9872 |
| Insulin - GLP-1R agonists | IgG - G2 | -0.6631 | 0.4257 | 0.1006 | 0.4616 |
| Insulin - GLP-1R agonists | IgG - S | -0.1447 | 0.5274 | 0.7671 | 0.8854 |
| Insulin - GLP-1R agonists | Plasma - AF | 0.2658 | 0.5366 | 0.5936 | 0.8081 |
| Insulin - GLP-1R agonists | Plasma - B | 0.0512 | 0.5221 | 0.9157 | 0.9509 |
| Insulin - GLP-1R agonists | Plasma - CF | 0.8629 | 0.4360 | 0.0396 | 0.4056 |
| Insulin - GLP-1R agonists | Plasma - G0 | 0.7412 | 0.4681 | 0.0953 | 0.4582 |
| Insulin - GLP-1R agonists | Plasma - G1 | 0.7053 | 0.3806 | 0.0530 | 0.4537 |
| Insulin - GLP-1R agonists | Plasma - G2 | -0.3464 | 0.5140 | 0.4687 | 0.7593 |
| Insulin - GLP-1R agonists | Plasma - G3 | -0.3809 | 0.4905 | 0.4046 | 0.7203 |
| Insulin - GLP-1R agonists | Plasma - G4 | -0.2405 | 0.5450 | 0.6343 | 0.8356 |
| Insulin - GLP-1R agonists | Plasma - HB | -0.4617 | 0.4934 | 0.3165 | 0.6837 |
| Insulin - GLP-1R agonists | Plasma - HM | -0.6498 | 0.5112 | 0.1768 | 0.5382 |
| Insulin - GLP-1R agonists | Plasma - LB | 0.6478 | 0.4704 | 0.1445 | 0.5382 |
| Insulin - GLP-1R agonists | Plasma - S0 | 0.8380 | 0.4209 | 0.0386 | 0.4056 |
| Insulin - GLP-1R agonists | Plasma - S1 | -0.2737 | 0.5485 | 0.5909 | 0.8081 |
| Insulin - GLP-1R agonists | Plasma - S2 | -0.3989 | 0.4988 | 0.3909 | 0.7203 |
| Insulin - GLP-1R agonists | Plasma - S3 | -0.4249 | 0.4993 | 0.3615 | 0.7203 |
| Insulin - GLP-1R agonists | Plasma - S4 | -0.2354 | 0.5494 | 0.6441 | 0.8356 |
| SGLT2 inhibitors - GLP-1R agonists | IgG - B | -0.1915 | 0.2332 | 0.3994 | 0.7203 |
| SGLT2 inhibitors - GLP-1R agonists | IgG - F | 0.0647 | 0.2528 | 0.7925 | 0.8854 |
| SGLT2 inhibitors - GLP-1R agonists | IgG - G0 | -0.0119 | 0.2296 | 0.9576 | 0.9757 |
| SGLT2 inhibitors - GLP-1R agonists | IgG - G1 | 0.1039 | 0.2431 | 0.6602 | 0.8356 |
| SGLT2 inhibitors - GLP-1R agonists | IgG - G2 | -0.1476 | 0.2253 | 0.5010 | 0.7698 |
| SGLT2 inhibitors - GLP-1R agonists | IgG - S | 0.1767 | 0.2470 | 0.4626 | 0.7570 |
| SGLT2 inhibitors - GLP-1R agonists | Plasma - AF | 0.0823 | 0.2496 | 0.7346 | 0.8837 |
| SGLT2 inhibitors - GLP-1R agonists | Plasma - B | 0.3384 | 0.2487 | 0.1645 | 0.5382 |
| SGLT2 inhibitors - GLP-1R agonists | Plasma - CF | 0.3153 | 0.2516 | 0.1999 | 0.5489 |
| SGLT2 inhibitors - GLP-1R agonists | Plasma - G0 | 0.1111 | 0.2463 | 0.6427 | 0.8356 |
| SGLT2 inhibitors - GLP-1R agonists | Plasma - G1 | 0.1892 | 0.2456 | 0.4291 | 0.7475 |
| SGLT2 inhibitors - GLP-1R agonists | Plasma - G2 | -0.0718 | 0.2579 | 0.7746 | 0.8854 |
| SGLT2 inhibitors - GLP-1R agonists | Plasma - G3 | -0.1079 | 0.2465 | 0.6529 | 0.8356 |
| SGLT2 inhibitors - GLP-1R agonists | Plasma - G4 | -0.0894 | 0.2565 | 0.7202 | 0.8837 |
| SGLT2 inhibitors - GLP-1R agonists | Plasma - HB | -0.1145 | 0.2492 | 0.6369 | 0.8356 |
| SGLT2 inhibitors - GLP-1R agonists | Plasma - HM | 0.1748 | 0.2546 | 0.4808 | 0.7658 |
| SGLT2 inhibitors - GLP-1R agonists | Plasma - LB | 0.0745 | 0.2493 | 0.7586 | 0.8854 |
| SGLT2 inhibitors - GLP-1R agonists | Plasma - S0 | 0.0408 | 0.2520 | 0.8678 | 0.9188 |
| SGLT2 inhibitors - GLP-1R agonists | Plasma - S1 | 0.2178 | 0.2562 | 0.3831 | 0.7203 |
| SGLT2 inhibitors - GLP-1R agonists | Plasma - S2 | -0.2413 | 0.2548 | 0.3316 | 0.6976 |
| SGLT2 inhibitors - GLP-1R agonists | Plasma - S3 | -0.0688 | 0.2483 | 0.7758 | 0.8854 |
| SGLT2 inhibitors - GLP-1R agonists | Plasma - S4 | -0.0450 | 0.2582 | 0.8577 | 0.9163 |
